# Assessment of Aerosol Persistence in ICUs via Low-cost Sensor Network and Zonal Models

**DOI:** 10.1101/2022.03.03.22271831

**Authors:** K Glenn, J He, R Rochlin, S Teng, JG Hecker, I Novosselov

## Abstract

The COVID-19 pandemic raised public awareness about airborne particulate matter (PM) due to the spread of infectious diseases via the respiratory route. The persistence of potentially infectious aerosols in public spaces and the spread of nosocomial infections in medical settings deserve careful investigation; however, a systematic approach characterizing the fate of aerosols in clinical environments has not been reported. This paper presents a methodology for mapping aerosol propagation using a low-cost PM sensor network in ICU and adjacent environments and the subsequent development of the data-driven zonal model. Mimicking aerosol generation by a patient, we generated trace NaCl aerosols and monitored their propagation in the environment. In positive (closed door) and neutral-pressure (open door) ICUs, up to 6% or 19%, respectively, of all PM escaped through the door gaps; however, the outside sensors did not register an aerosol spike in negative-pressure ICUs. The K-means clustering analysis of temporospatial aerosol concentration data suggests that ICU can be represented by three distinct zones: (1) near the aerosol source, (2) room periphery, and (3) outside the room. The data suggests two-phase plume behavior: dispersion of the original aerosol spike throughout the room, followed by an evacuation phase where “well-mixed” aerosol concentration decayed uniformly. Decay rates were calculated for positive, neutral, and negative pressure operations, with negative-pressure rooms clearing out nearly twice as fast. These decay trends closely followed the air exchange rates. This research demonstrates the methodology for aerosol monitoring in medical settings. This study is limited by a relatively small data set and is specific to single-occupancy ICU rooms. Future work needs to evaluate medical settings with high risks of infectious disease transmission.

## 1 Introduction

Nosocomial infections can occur during the patient’s stay in the hospital or after visits to medical facilities. Many of these diseases can be transmitted through the respiratory route, associated with exposure to airborne microparticles generated by coughing, sneezing, talking, aerosol-generating medical procedures, or particles resuspended from surfaces. These nosocomial aerosols may contain infectious biological organisms, e.g., bacteria, viruses, and fungi. Exposure to these aerosols presents a risk to patients, especially compromised individuals, and to medical staff ^1^. The Extended Prevalence of Infection in Intensive Care report suggests that the number of infected patients in intensive care Units (ICUs) can be greater than 51%.^2^ There is a need to investigate the transmission risk and develop mitigation strategies due to increased hospital stays and increased antimicrobial resistance.

Since early 2020, exposure to the SARS-CoV-2 virus has been a concern for patients and hospital staff. SARS-CoV-2 can be spread by inhaling aerosols containing viable virions like other respiratory diseases. ^3, 4^ Hospital environments are particularly susceptible to the spread of infection, and evidence-driven approaches can aid in combatting the spread of airborne diseases.^5^ Surfaces and air samples from ICUs with COVID-19 patients showed that while all surface samples tested negative, air samples remained positive, implying that SARS-CoV-2 shed and persisted as aerosol days after a patient tested negative. ^6^ To minimize airborne transmission in densely occupied spaces, upgrades to the ventilation systems have been suggested. ^7^ While there have been some efforts to model and measure aerosol movements within hospital environments ^8^, there has been a lack of reports on the systematic characterization of aerosols in hospital environments. Negative pressure ICUs are currently used for controlling the spread of infectious aerosols. It has been suggested that aerosols persist in negative pressure ICUs for 20 - 30 minutes, and healthcare providers should wear fitted respirator masks and other personal protective equipment (PPE). ^4, 9^ Without the characterization of aerosol distribution, these recommendations remain speculative and are not general enough to apply to all ICUs or other medical settings.

Computational fluid dynamic (CFD) simulations have been used for characterizing indoor air aerosol dynamics. ^10^ However, uncertainties in boundary conditions (flow rates, exact locations of air vents and intakes), modeling of particle turbulence interaction^11^ and computation cost and setup times present challenges in implementing CFD as an effective tool for heating, ventilation, and conditioning (HVAC) systems design and air quality control strategy. Adequately resolving large volumes by transient CFD simulations requires large numerical grids, leading to costly simulations, and validation of the CFD models against time- and space-resolved data is needed. For example, Buchanan and Dunn-Rankin reported that aerosol distribution patterns in operating rooms (ORs) highly depend on the characteristics of each room. ^12^ CFD simulations of flow in ICU have been used to gain insight into the flow patterns ^13^, specifically relevant are the recent studies that addressed the transport of potentially infectious aerosols. ^14, 15^ These time-consuming CFD computations can benefit from validation using rapid spatiotemporal aerosol concentration measurements. E.g., data in this work using sensors were available in real-time, and each experiment took 5 - 10 minutes.

An approach that uses empirical data can inform targeted aerosol mitigation strategies and HVAC system optimization. Tracer methods are currently used in indoor air quality investigations; these typically use tracer gases rather than aerosols.^16^ For example, both the American Society of Heating, Refrigerating and Air-Conditioning Engineers (ASHRAE) and the American Society for Testing and Materials (ASTM) International have standardized test procedures. ^17^ The use of tracer gases is a common approach for acquiring quantitative data on ventilation rates; however, it may not fully represent aerosol behavior. Recently several researchers utilized tracer aerosols to evaluate air quality and aerosol propagation in indoor environments. ^18^ Motivated by the need to study the risk of infectious disease transmission, Edwards et al. used networked PM counters to study aerosol spread on public busses using nebulized NaCl particles ^19^, and Makhsous et al. used a similar method to study aerosol transport in dental offices. ^20^ In our study, the nebulizer produced a visible water vapor plume that persisted for ∼ 0.5 m from the outlet; the droplets evaporated, yielding dry NaCl particles. Gravity effects such as droplet settling ^21^ were not evaluated, as only smaller aerosols (< 3 µm) were generated.

Recent advancements in low-cost particulate matter (PM) sensors led to their extensive use in various applications, such as air quality (AQ) in indoor environments ^22^ and outdoor ^23^, including large-scale deployments ^24, 25^ by academic researchers and citizen scientists. Optical PM sensors rely on elastic light scattering, providing size-resolved PM concentrations in the 0.3 – 10.0 μm range. The low-cost sensor measurements often suffer from sensor-to-sensor variability due to a lack of quality control and differences between individual components.^26^ The scattering light intensity depends on particle size, morphology, complex index of refraction (CRI), and sensor geometry. ^27^ CRI sensitivity can be addressed by optimizing the design to measure scattered light at multiple angles simultaneously or by employing dual-wavelength techniques. ^28^ However, these solutions are complex and involve expensive components that are not suitable for compact, low-cost devices.^20^ One of the main limitations is that low-cost optical sensors act as integrating nephelometers, not optical single-particle counters; they measure total light scattering and cannot provide accurate measurements of all concentration and particle size bins ^29^; thus, sensor calibration and data quality control are required.

Various studies have evaluated the performance of low-cost PM sensors in laboratory and field settings.^30^ These reports show that low-cost sensors yield usable data when calibrated against research-grade reference instruments.^25, 31, 32^ The sensor networks have the potential to provide high spatial and temporal resolution, identifying pollution sources and hotspots, which in turn can lead to the development of better exposure assessment and intervention strategies for susceptible individuals.^33^ Data can be fitted to a regression model when analyzing large data sets from such networks. Still, for a large number of sensors and sequentially collected data, conventional techniques may not suffice. Problems with larger dimensions require sequential (or recursive) estimation approaches.^34^ An essential factor in the data analysis to simplify or condense the data is to aggregate data from sensors providing that the specifics of the data (such as the locations of the sensor nodes) are not left out.^35^ All sensor networks must also have time synchronization.

There is no single technique for analyzing distributed sensor network data; among them are K-means clustering algorithms, which have been used in several wireless sensor networks.^36, 37^ The K-means algorithm is efficient at distributed sensor clustering (the method used in this paper), allowing every node to inform clustering decisions instead of choosing a ‘head’ node for each cluster. Clustering algorithms are practical in pattern recognition and statistical analysis. ^37^

In summary, there is a need to characterize aerosol persistence in the ICU and other medical settings. There is a lack of space - and time-resolved data on the persistence of aerosols in clinical environments. This paper reports a methodology for obtaining real-time space-resolved data on the persistence of aerosols in ICUs using a low-cost sensor network. The data from multiple experiments were used in the K-means regression analysis yielding zonal models for aerosol persistence in ICUs. The zonal model is used to evaluate the difference between positive, neutral, and negative pressure rooms in terms of aerosol decay rates, distribution patterns, and aerosol exfiltration from the ICU. This approach provides a predictive tool for rapidly assessing aerosol persistence in real-world settings.

## 2 Methods

### 2.1 Intensive Care Units

We present a low-cost approach for evaluating the effectiveness of ventilation systems in clinical facilities to minimize aerosol dispersal into shared spaces. Experiments were conducted in four ICUs at the University of Washington Medical Center (UWMC). The data capture the spatial and temporal distribution of aerosols in ICUs and the immediate vicinity. We analyzed and modeled the dispersion and exfiltration of aerosols generated in the ICUs, gaining insight into the potential for exposure inside and outside the rooms under real-world conditions. Four ICU rooms were studied, and two (ICU 1 and ICU 3) were tested in positive and negative pressure configurations. Table 1 shows room dimensions and the air exchange rates (ACR) per hour for each ICU provided by the UWMC facilities operation and maintenance department. The ICUs with negative pressure capability show higher ACR in the negative mode (by 60 – 115 %). In the negative mode, the flow rate depended on the performance of the blower in the specific room.

**Table 1:**
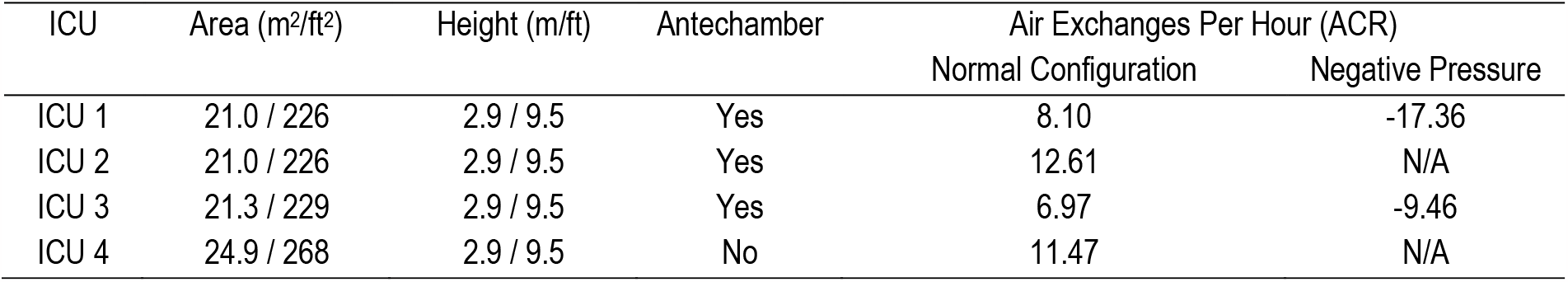
Air Exchanges per Hour for each ICU in normal and negative configurations. The UWMC maintenance department provided the ACR rates.

Airflow velocities for each outlet and the door gap at the bottom of the door were measured using a hot-wire anemometer (HT-9830, Hi-Tech Instruments, Pomona, CA). Dimensions of the vents and door gaps were measured; they were used to calculate the fraction of air flow rate escaping from the room. The procedure was performed for each experiment and averaged between three replicates to determine the confidence intervals.

### 2.2 Low-cost Particulate Matter Monitor

Time and space-resolved 3D aerosol monitoring require compact devices with accurate particle sizing and a fast sampling rate.^38^ This study utilized custom PM monitors that incorporate PM sensors, humidity and temperature sensors, Wi-Fi, and a communication module for data transfer; more detailed information on the monitor design can be found in ref ^39^. Briefly, the data can be transmitted in real-time to the database using a Wi-Fi or cellular connection; the secure digital (SD) card is used for onboard data backup. The data acquisition rate was set to 10 seconds. The schematic and photograph of the monitor are shown in Supplementary Figure 1. Each unit incorporates Plantower PMS A003 (Plantower, Beijing Ereach Technology Co., Ltd, China). A sensor’s photodiode positioned normal to the excitation beam measures the light scattered by the particles in the optical volume. The scattering light intensity is then converted to a voltage signal to estimate PM number density and mass concentration. The PMS provides particle counts in 6 size bins in the optical diameter range of 0.3-10 µm range and mass concentration (µg/m^3^) for PM_1_, PM_2.5_, and PM_10_. Here, original equipment manufacturer (OEM) calibration in a standard setting was used.

The performance of these sensors was assessed during environmental monitoring campaign measuring indoor and outdoor exposure to wildfire smoke. ^40^ However, for NaCl aerosols plume tracking, it is most appropriate to evaluate sensor-to-sensor repeatability with the same particle type. Our previous PMS calibration experiments in the well-mixed aerosols test chamber ^41 42^, showed that sensor-to-sensor repeatability for NaCl aerosols was within 15%. ^31^ The NaCl particles were generated by nebulizing the aqueous solution of NaCl 10% wt. ^43^ The total particle number concentration in the test chamber was maintained below 1000 #/cm^3^. In these tests, PMS units were installed on a custom printed circuit board connected to an Arduino Nano microcontroller. The controller simultaneously collected data from the PMS sensors, relative humidity (RH), and temperature sensors. An aerodynamic particle sizer (APS, TSI, model 3321) was used as a reference instrument to measure particle concentration and size distribution. The PMS sensors were calibrated against the APS concentration and PM size distribution. ^31^

Before this hospital campaign, which included the ICU and OR measurements reported by Hecker et al. ^44^, the sensors were tested in the aerosol chamber against NaCl particles. Here, we report the total number concentration, i.e., all particles with an optical diameter > 0.3 µm (all size bins combined). In a typical experiment, the nebulizer generated NaCl particles < 3 μm, measured by the APS, with a median aerodynamic diameter of 0.86 µm. This size fraction stays suspended in the air for an extended time due to its low settling velocities, and it is a suitable surrogate for long-lived aerosols that may contain SARS-CoV-2 or other infectious agents. ^19, 45^ Additionally, spot checks were performed during ICU experiments where sensors in the network grid were compared to a “reference” PMS sensor that was moved between different grid locations periodically, see Figure 1.

**Figure 1:**
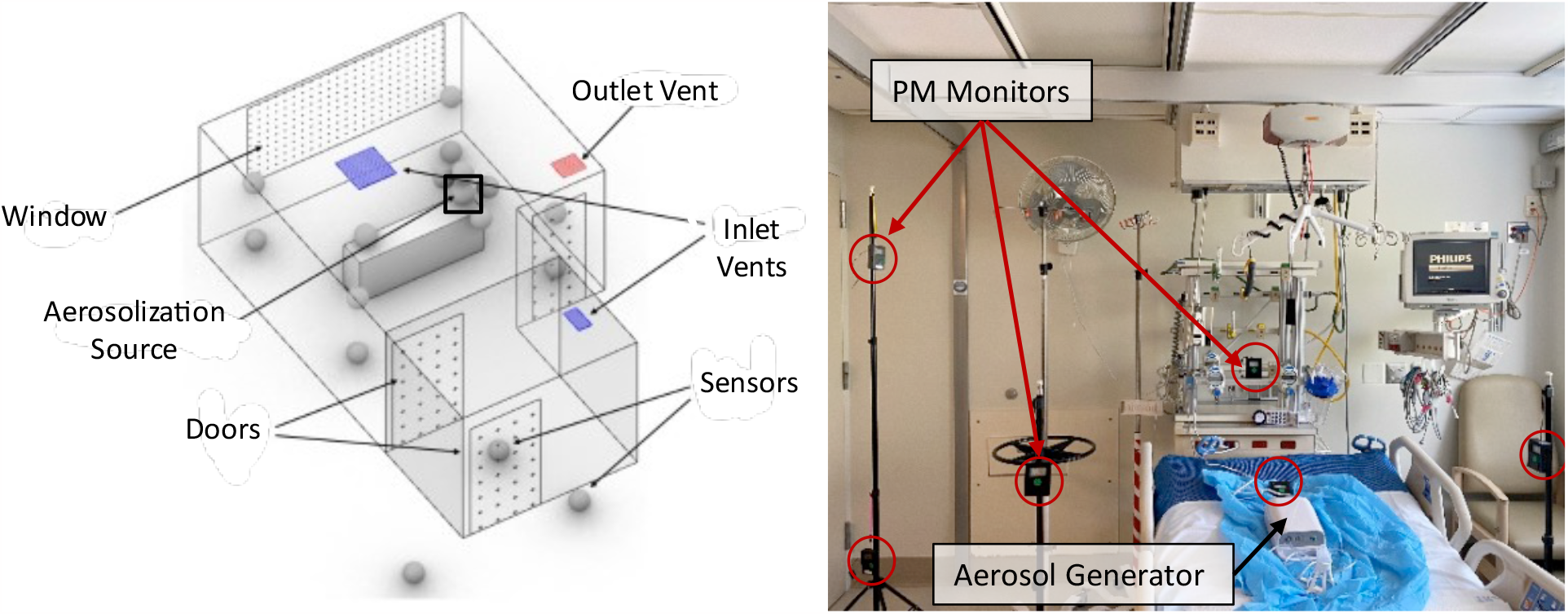
Distributed low-cost sensor network gathers time and space-resolved aerosol information. The data is transmitted to a cloud server, displayed in near real-time, and saved on the SD card. Left: Schematic of typical room layout and sensor positions. The black square represents the location of the particle generation source. Right: Photograph of the sensors in the ICU.

### 2.3 Sensor Network

To capture the spatial distribution, we deployed a sensor network consisting of 16 - 18 aerosol monitors in a predetermined grid inside the ICUs and the common areas outside the rooms, shown in Figure 1. Each monitor in the network is shown as a sphere. The 2-3 hallway sensors are located near the door gap and another in the hallway about 10-20 feet from the door. At least one sensor was placed in the antechamber in the negative pressure ICUs (see Figure 1). The monitors were placed at three different heights – about 0.2 m, 1.0 m, and 2 m (each kept ∼ 1 – 2 m apart for an evenly spaced grid). Aerosol particles were generated at the head of the bed, mimicking aerosol release during the patient’s coughing, sneezing, or talking.

### 2.4 Experimental Procedure

During the experiments, the ICUs were unoccupied except for the researchers who tuned the nebulizer ON and OFF and a researcher who monitored the periphery sensor(s) reading to indicate the end of the data collection. Once the particle concentration returned to the baseline level, the next replicate experiment was started. The sensors’ and nebulizer locations did not vary between the replicate experiment or when the pressure conditions were switched. NaCl particles were generated at the patient’s head’s location, and the sensor monitored the aerosol concentration in real time until all PM measurements returned to the background level. Particles were generated for 60 sec at a steady rate by nebulizing 1% NaCl solution using the MADA Up-Mist™ medication nebulizer (MADA Products, Carlstadt, NJ, USA), generating polydisperse particles in 10 nm – 3 µm size range as measured by Nanoscan (TSI 3910) and APS 3321 in the aerosol chamber calibration experiments which agrees with the previous work.^46^ Once aerosolized, particle behavior is governed by aerodynamic properties, and the chemical composition does not affect their persistence in the environment. The 60-second aerosolization produced enough particles for plume tracking (100x - 1000x higher than the background concentrations) but not enough to saturate the sensors and trip ICU smoke detectors, see Figure 2. Our previous study in ORs ^44^ tested 300 sec aerosolization; however, the sensors in the vicinity of the nebulizer became saturated. The 60-sec nebulization produced an initial spike, following which the particle concentration decayed as the air was exchanged via an HVAC system (through air outlets) or escaped through gaps under the doorway or the open door. Experiments were terminated when the aerosol concentration in the room reached the background level, < 1 #/cm^3^, based on the total particle count. A short nebulization is also beneficial for getting time-resolved data, as a longer nebulization resulted in simultaneous particle generation and evacuation, thereby confounding the analysis.

**Figure 2:**
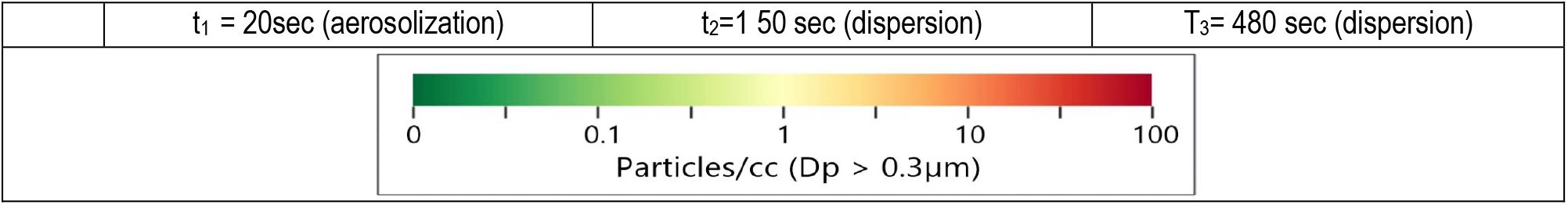
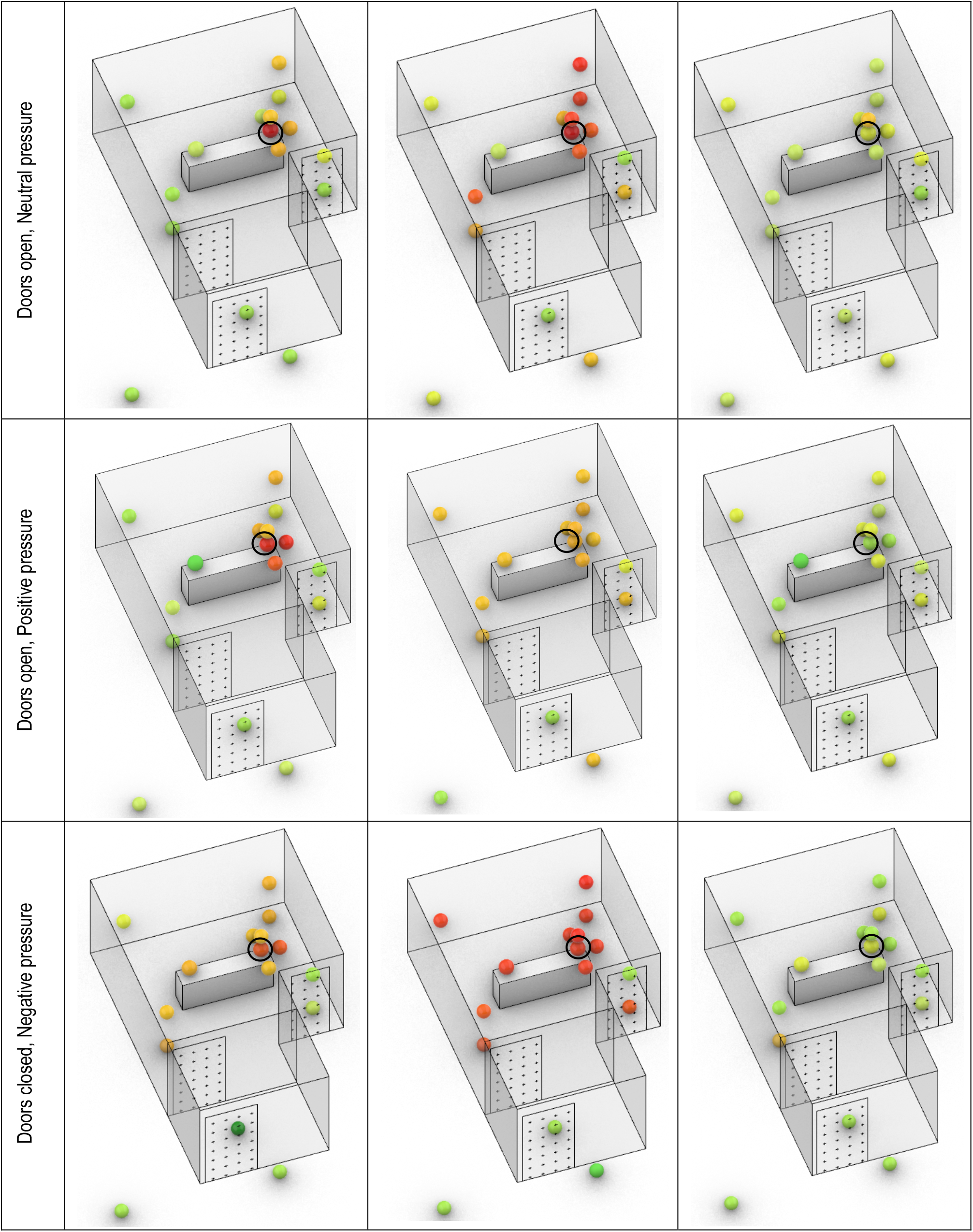
3D maps from ICU 1 with positive pressure (door closed), neutral pressure (door open), and negative pressure. NaCl particles are nebulized for 60 seconds at the location of the patient’s head. Back hallway sensors are located 10-20 feet from the door. The black circle indicates the aerosol source site.

### 2.5 Data Collection and Analysis

For each experiment, the PM data timestamps were first aligned to the start of the nebulization, which was used as the datum. The sensor readings were averaged in 10-second epochs for each sensor; these data were used to create a time-resolved 3D map of aerosol distribution. Each sensor’s particle concentration data with timestamps were saved locally to the secure digital (SD) card. Triplicate measurements were made for each condition: doors open (neutral pressure), closed (positive pressure), and negative pressure. Based on the spatiotemporal information, the sensors were grouped into zones using K-means clustering. ^47^ The algorithm iteratively finds a globally optimal data partition into a specified number of clusters. ^48^ A clustering algorithm assigns sensors with similar behavior, such as the ‘peak’ value and when the peak occurred during the experiment for each sensor. Since the algorithm uses unsupervised learning, the only user-inputted data is the number of zones desired and the actual data. The K-means optimization was implemented using a custom Python code for each experiment and each ICU. In the initial analysis, the number of zones varied; after initial trials, the teams settled on using three zones to streamline the analysis.

## 3 Results and Discussion

### 3.1 3D Data of Aerosol Distribution

The data for each experiment are plotted as a 3D map at given timestamps (Figure 2), as a 2D map (Supplementary Figure 3), and as a decay plot vs. experimental time (Figure 3). Figure 2 shows a typical 3D-resolved aerosol distribution map during the aerosolization and dispersion phases in ICU 1 for positive, neutral, and negative pressure operations. All experiments show the initial concentration spike during aerosolization at the head of the bed and uniform spreading in the dispersion phase. Both positive pressure and neutral pressure conditions showed similar trends. After the initial PM spike during the aerosolization, at ∼90 – 120 sec, the aerosol concentration in the room reaches a well-mixed state, i.e., sensors near the aerosolization source and the periphery sensors have similar readings. At the same experimental time, the particle plume reaches the outer door of the ICU and begins escaping the room; the particle concentration outside the ICU door reaches its maximum. The particle plume becomes diluted as it enters the common area. The monitors did not register the particles emitted from the room 10-20 feet from the ICU door.

**Figure 3:**
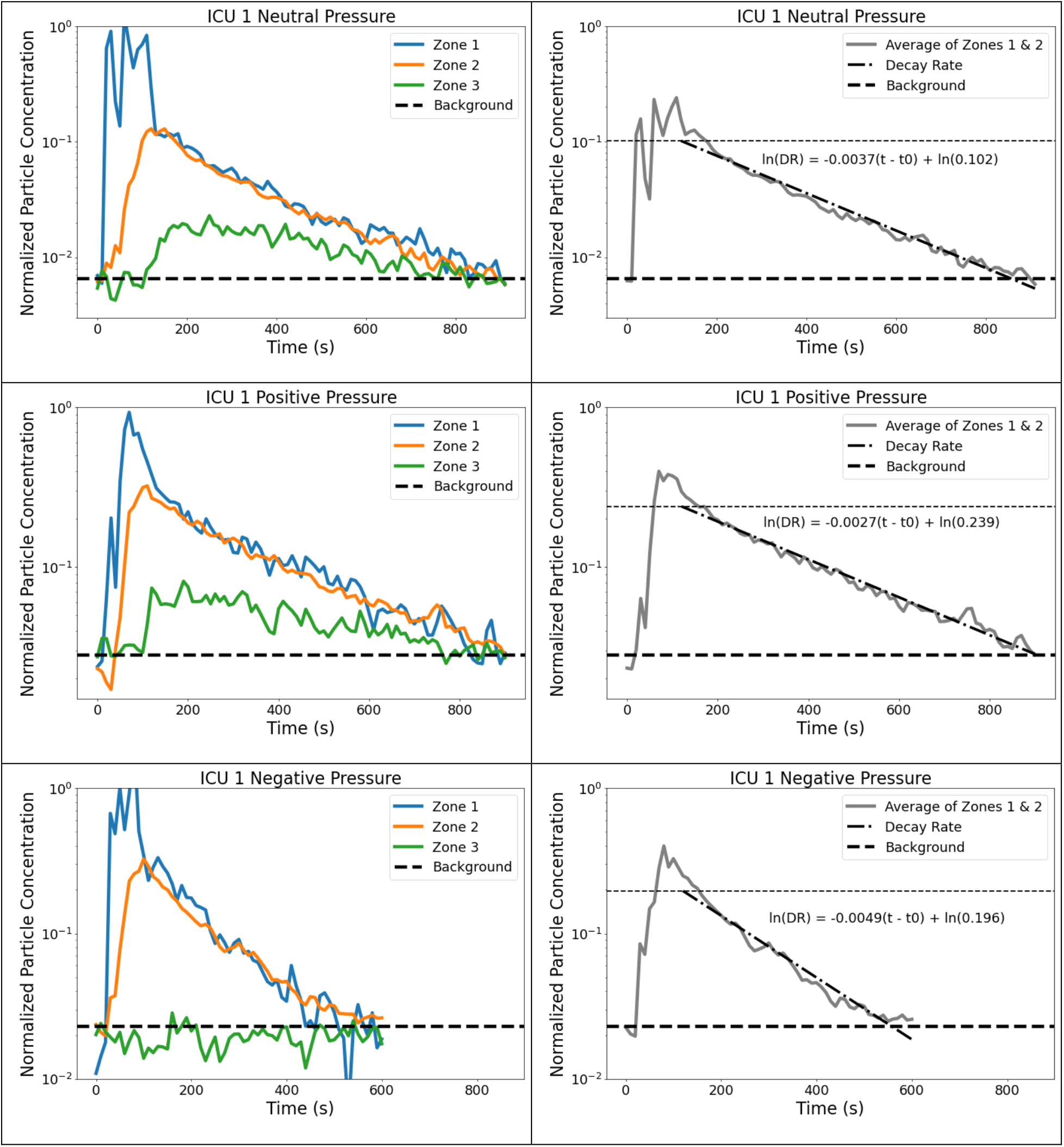
Normalized time data from ICU 1 showing aerosol concentration for each zone. Left: Zones compared to background levels. Right: Normalized average data with decay rate starting 60s after the end of nebulization. The higher horizontal line represents the value of A, or the normalized value of aerosol at which the room becomes well-mixed.

The aerosol concentration in the room decays as it becomes diluted by HVAC-filtered air and exhausted via the vent in the room. The well-mixed condition suggests that air currents in the room produced by HVAC effectively mix the trace particle before evacuating the aerosols completely. This observation can be contrasted with a study by Hecker et al. in ORs utilizing a similar experimental design.^44^ The larger ORs did not exhibit well-mixed conditions, and the aerosol distribution could be clearly defined between different OR zones.

The concentration spike near the aerosolization source was the lowest with negative pressure as the aerosol was effectively mixed inside the room. This is likely due to the higher ACR, resulting in greater air velocities (greater turbulence levels) in the room that enhance mixing. The high ACR resulted in a faster return to the baseline level of ∼ 500 sec than ∼ 800 sec in the positive and neutral pressure cases. Even more importantly, the outside sensors did not register any spikes in aerosol concentration for negative pressure rooms, indicating that the particles generated in the ICU remained in the room. This observation confirmed the efficacy of negative pressure ICU for controlling infectious diseases.

### 3.2 Zonal Analysis

Based on the spatiotemporal trends, the environment can be split into zones where the aerosol concentrations and persistent trends are similar. While these zones could be assigned based on physical location, the data-driven approach can provide insight into aerosol behavior and persistence without *a priori* assumptions and can account for PM sources, sinks, and airflow patterns. The zonal maps based on K-means clustering for ICU 1 are shown in Supplementary Figure 2. Similar to the 3D mapping analysis, the sensor data from three replicate experiments were averaged, and three scenarios (positive, neutral, and negative pressure) were evaluated independently for each ICU. Zone 1 sensors had the largest and the earliest spike in PM concentration. Zone 2 sensors spiked later, and their PM levels were consistent. Zone 3 sensors had a lower and delayed PM spike or did not rise above the background levels in the negative pressure ICU. Note that zone assignment is a function of threshold level (variance between the sensors) set by the user, and the zone assignment can vary, depending on these settings. For example, in ICU 1, the algorithm assigned a different split between Zone 1, Zone 2, and Zone 3. The positive and neutral pressure zone assignments were nearly the same. However, for the negative pressure cases, the Zones were significantly different, e.g., both sensors at the head of the bed were assigned to Zone 2, likely due to the strong suction vent near the head of the bed. SI Fig. 2 shows that the airflow setting affects the zonal assignment. Thus, for further analysis, a zonal map for each ICU was fixed based on the positive pressure ICU data, and each ICU was analyzed separately.

Figure 3 (left) shows the time series of the zonal PM concentrations in ICU 1. Due to variability in initial PM concentration (#/cm^3^), we normalized the data by dividing all the values by the peak value of that experiment. The normalization is necessary to compare the data across all rooms, and all conditions since the initial particle concentration may vary depending on the aerosol generation, nebulizer position, flow currents in the room, room geometry, and other factors. Consequently, all the decay rates are plotted on the same scale with the maximum value of unity. For the initial PM loading, the aerosol concentration returned to the background level (∼1 #/cm^3^) in positive and neutral pressure ICU at ∼ 800 sec and negative pressure ICU at ∼ 500 sec. Individual zone behavior can be generalized as follows:

- Zone 1 -- the concentration spikes immediately with particle generation and then equilibrates to the average room concentration within 30 – 60 sec after aerosolization is stopped.
- Zone 2 -- PM concentration rises within 10 – 30 sec and reaches the maximum when the initial peak in Zone 1 is reduced but before the aerosol is diluted by the introduction of HVAC-filtered air. This indicates that generated aerosol at a point source is first dispersed through the room before it is evacuated by the HVAC system -- the room reaches well-mixed condition.
- Zone 3 -- in the case with positive or neutral pressure, the PM levels are lower than in the room as the aerosol plume exits the ICU through the gaps around the door or the open door. The PM levels are at the background level for negative pressure as the air enters the room from outside.

Though both internal mixing and removal of PM from the room co-occur, the timescale for these processes in the ICU is different by order of magnitude. The dispersion of the initial spike from Zone 1 to the rest of the room happens over ∼ 30 – 60 sec, while the aerosol evacuation (return to the background) takes ∼ 500 – 800 sec. This separation of time scale suggests that two phases can be identified: (i) dispersion phase and (ii) evacuation phase. The dispersion phase is characterized by the spatial redistribution of the initial PM spike to the room’s periphery (from Zone 1 to Zone 2). After the aerosolization is stopped, the PM concentration in Zone 1 rapidly returns to the room’s average concentration level due to air mixing. During the evacuation phase, the aerosol from a well-mixed room is removed by the HVAC system or exfiltration through the doors, door gaps, or other openings. The air exchange rate and room condition (i.e., door open or closed, the pressure differential between the room and the outside) governs how fast the room returns to the background PM concentration. In positive and neutral pressure conditions, the aerosols may escape from the room (from Zone 2 to Zone 3); in the negative pressure operation, the air from the adjacent environment infiltrates into the ICU following the pressure differential (from Zone 3 to Zone 2). Additional studies are necessary to determine if the distinction between the phases can be generalized to other settings; the indoor settings, ventilation rates, type of aerosol generation sources, filtration strategies, and other variables are likely to play a role in the duration and overlap of these phases. The sensor network approach can obtain the necessary temporal and spatial resolution for these analyses.

### 3.3 Aerosol Exfiltration Rates

After the well-mixed condition is achieved (t ∼ 90 – 120 sec or 30 – 60 sec after aerosolization stopped), the aerosol concentration decays uniformly, allowing characterization of the decay rate directly by observing the average particle concentration in the room. During the evacuation phase, the particle concentration was similar for all sensors within the room; thus, we averaged the data of all sensors in the ICU (Zone 1 and Zone 2). Figure 3 shows that the concentration (*P*, #/cm^3^) decays linearly when plotted on the log scale against the experimental time. Thus, particle concentration can be fitted to expression in the form *P = P*_*0*_ *exp (b t*)*, where *P*_*0*_ is the concentration at *t = t*_*0*_ when the well-mixed state is reached, *b* -- the decay rate and *t* = (t – t*_*0*_) is the experimental time from the well-mixed condition. Here, in all cases, *t*_*0*_ was set to a more conservative value of 60 sec after the end of aerosolization. Figure 3 shows the aerosol concentration decay in ICU 1 under positive, neutral, and negative pressure operations. The expressions for all ICUs are presented in Table 2. The positive and neutral pressure operations have similar decay rates and clear-out times for a given ICU due to consistent experimental procedures and similar initial aerosol spikes. The decay rates are calculated from the sensors’ data inside the room and do not include the sensors in Zone 3.

**Table 2:** Decay rate and exfiltration analysis; the first column for each scenario shows the time it takes for the room to return to background levels, and the second column shows the decay rates.

|  | Neutral Pressure |  |  | Positive Pressure |  |  | Negative Pressure |  |  |
| --- | --- | --- | --- | --- | --- | --- | --- | --- | --- |
|  | Time to Clear (sec) | 90% Thres hold (sec) | Decay Rate (DR) | Time to Clear (sec) | 90% Thres hold (sec) | Decay Rate (DR) | Time to Clear (sec) | 90% Thres hold (sec) | Decay Rate (DR) |
| ICU 1 | 830 | 191 | $0.070\exp(-0.0036t^*)$ | 830 | 178 | $0.057\exp(-0.0033t^*)$ | 530 | 191 | $0.167\exp(-0.0076t^*)$ |
| ICU 2 | 502 | 361 | $0.566\exp(-0.0089t^*)$ | 565 | 296 | $0.392\exp(-0.0088t^*)$ | N/A | N/A | N/A |
| ICU 3 | 821 | 224 | $0.102\exp(-0.0037t^*)$ | 830 | 265 | $0.239\exp(-0.0027t^*)$ | 450 | 201 | $0.196\exp(-0.0049t^*)$ |
| ICU 4 | 465 | 160 | $0.042\exp(-0.0068t^*)$ | N/A | N/A | N/A | N/A | N/A | N/A |

Table 2 shows the time to reach a 90% reduction and the time to return to background PM concentration for each scenario in each ICU. Note that in the negative pressure scenario, the particle-laden air was aspirated into the ICU by HVAC, and in the neutral and positive pressure experiments, the aerosol was mixed through the door opening. Thus, for consistency, the sensors located in the hallway (10 – 20 feet away from ICU) were used for background measurements.

Table 2 shows the empirical decay rate expression (*DR*) to estimate the relative concentration of aerosol after the room reaches well-mixed conditions. The ACR (see Table 1) correlates well with the decay rates for ICU. For example, a comparison of the ICU1 and ICU3 (with the nearly identical layout) shows that, in the negative mode, the ICU1/ICU3 ACR ratio was 1.83, and the decay rate exponent ratio was 1.6. The trends hold for positive mode for ICUs of similar geometries, suggesting that measuring aerosol concentration allows for direct comparison of the efficacy of HVAC performance in ICU. Other environments, such as ORs and emergency rooms, may require more detailed measurements due to nonuniform spatiotemporal aerosol concentrations.^44^

Table 3 estimates the fraction of aerosolized microparticles evacuated by the HVAC in the ICU vs. the fraction exfiltrated from the room to the common areas. The analysis was performed for well-mixed conditions by comparing the data from the inside (Zones 1 and 2) and outside the room (Zone 3). First, the PM concentration recorded by each sensor (# / cc) was corrected to the coverage area (volume) for the particular location, which is possible for the evenly distributed sensor grid and well-mixed PM concentration. Then the particle count from each sensor was added to determine the instantaneous particle count in each zone. In the evacuation phase, the particle count in each zone was integrated in time for the entire experiment, allowing us to determine the fraction of aerosolized particles that stayed in the room and were evacuated by HVAC vs. exfiltrated to the shared area.

**Table 3:** Percentage of aerosols dispersed to each location (inside the room, under the doorway, and at the nursing station) after aerosolization.

| ICU | Negative Pressure |  |  | Positive Pressure |  |  | Neutral Pressure |  |  |
| --- | --- | --- | --- | --- | --- | --- | --- | --- | --- |
|  | Inside (%) | Doorway (%) | Nurse Station (%) | Inside (%) | Doorway (%) | Nurse Station (%) | Inside (%) | Doorway (%) | Nurse Station (%) |
| ICU 1 | 103.4 ± 0.8 | -2.5 ± 0.5 | -0.9 ± 0.3 | 101.3 ± 0.9 | -0.3 ± 0.9 | -1.0 ± 0.0 | 97.5 ± 2.0 | 2.9 ± 1.8 | -0.5 ± 0.3 |
| ICU 2 | N/A | N/A | N/A | 94.1 ± 1.9 | 4.3 ± 1.4 | 1.6 ± 0.6 | 86.3 ± 4.2 | 14.2 ± 1.4 | 0.6 ± 0.3 |
| ICU 3 | 101.8 ± 1.3 | -1.2 ± 0.9 | -0.5 ± 0.4 | 97.5 ± 2.7 | 6.0 ± 0.1 | -3.5 ± 2.6 | 95.1 ± 1.5 | 5.6 ± 1.8 | -0.8 ± 0.5 |
| ICU 4 | N/A | N/A | N/A | N/A | N/A | N/A | 79.8 ± 2.1 | 18.5 ± 1.3 | 1.7 ± 1.0 |

The following calculations were performed to find the fraction of particles leaving the room. (1) Volumetric flow rates (*Q*_*i*_) were calculated for each of the outlets by multiplying average velocity (*U*_*aver, i*_) measurements and area (*A*_*i*_), such as *Q*_*i*_ *= A*_*i*_ *U* _*aver, i*_, where index “*i* “denotes specific outlet. (2) The flow fraction for each outlet (*FF*_*i*_) is then calculated as *FF*_*i*_ *= Q*_*i*_ / *∑ Q*_*i*_. (3) Finally, to calculate the aerosol fraction for each outlet, the aerosol concentration measured in the vicinity of the outlet was integrated over time of the experiment and multiplied by the flow fraction of each outlet. Such as to calculate the aerosol fraction leaving the room through the door gap, the averaged data from two sensors positioned at the door gap were used.

The authors recognize that several uncertainties complicate the analysis, such as uneven sensor distribution and nonuniform PM concentration within the Zones; thus, this analysis provides the trends in the aerosol fate assessment and not exact values.

The zone immediately outside the ICU in the neutral and positive pressure scenarios shows the presence of fugitive aerosols. The open-door cases show the highest amount of exfiltrated aerosols from the ICU, ∼ 2.4% - 20.2%, and the close-door positive pressure case, ∼ 0% - 5.9%. In the experiments with closed-doors negative pressure, no fugitive aerosols were detected. Note that some values in the table have a negative percentage because of the varied background concentration in the hallway. If the concentration dips below this base value, it shows a negative percentage, and another zone might show a percentage of over 100% to account for that. The nursing stations outside the ICU (10 – 20 ft away from the ICU door) did not show a spike in the PM levels due to the dilution of the trace aerosol and high background levels. Using a more specific tracer may improve the analysis of fugitive aerosols. The zonal model suggests the surveillance of aerosols in the hospital may not require a dense measurement grid, and for ICU, 1-3 sensors may be sufficient. While tracer particles were used for plume tracking and the aerosol size distribution was not used in the analysis, in the surveillance applications, the particle size data could be instrumental for (i) determining the source of the particles and estimation of their fate (settling or removed by HVAC or other filtration units). Additional aerosol plume tracing experiments could be beneficial to generalize the aerosol propagation trend at UWMC; however, this study was performed in the working hospital during the Covid-19 pandemic, and extensive characterization of the ICU suite was challenging.

### 3.4 Comparison with CFD simulation

During the Covid -19 pandemic, there have been an increasing number of modeling papers investigating the flow patterns and aerosol distribution in medical facilities. The review of these studies is beyond the scope of this work; however, we can use our work to inform the need for future simulations and to potentially validate the CFD. There has been great enthusiasm for using CFD methods for analyzing aerosol distribution in indoor environments; ambiguity in the boundary conditions, computational methods, and the coupling between aerosol behavior and fluid motion requires additional validation before the CFD methods can be widely utilized.^11^ Computational and setup times often are significantly higher than direct measurement approaches presented in this work. Buchanan and Dunn-Rankin, performed CFD of flow distribution in an operating room, incorporating room dimensions, temperature, and personnel movement. ^12^ They concluded that aerosol distribution patterns highly depend on the characteristics of each room. Hecker et al. reported the aerosol distribution in the OR using a sensor network, confirming that the room layout significantly affects the aerosol distribution.^44^ E.g., in larger (and more modern) ORs, the aerosol plume did not reach the room perimeter due to an effective air handling scheme. The smaller rooms behaved similarly to the current ICU study. In all cases, the positive pressure environment resulted in significant aerosol leakage from the room. Another example of sensor network measurement in a medical facility (dental offices) was presented by Makhsous et al.^20^ The authors showed that the aerosol in the facilities with open floor plans, the aerosol generated from dental procedures, and simulated procedures with NaCl aerosols did not spread significantly passed the operatory but affected the dentist or technician.

Though this work presents the measured aerosol distribution in a smaller ICU for the first time, the results can be compared with the published CFD simulations. One of the benefits of CFD simulations is the ability to study the persistence of large particles and droplets and potentially the droplet evaporation on aerosol distribution. Wang et al. showed that fine particles are greatly affected by the HVAC setup, and large particles and droplets are mainly removed by surface deposition. ^14^ The authors also compared 3 types of ventilation patterns and concluded that the flow patterns greatly influenced the removal of particles; however, the temporal information about the decay rates was not provided, likely due to the challenges and computational cost of performing transient CFD simulations. Prajapati et al. showed the aerosol distribution in ICU following a 2-zone model, as was experimentally shown in our work. ^49^ Anghel et al. commented on the importance of the position for inlet and outlet grids and flow rate, leading to uneven aerosol distribution through the room. The authors suggest that the onset of local turbulences could influence plume propagation. ^50^ Our ICU and early OR measurements ^44^ confirm this observation, especially for larger rooms (as was modeled in the paper) or early in the particle generation cycle.

Crawford et al. performed transient Lattice Boltzmann Method (LBM) simulations to study the movement of airborne particles produced by a cough in a negative-pressure ICU. The CFD airflow patterns were compared with the Schlieren experiments. The results suggest that bed orientation and additional portable unit positioning can increase aerosol extraction by 40 % after 45 sec. ^15^. The application of LBM to study flow patterns is attractive due to potentially faster run time and the ability to run transient simulations more effectively; it is well suited for multiphysics problems ^51^, including stochastic flow description ^52^, and multiphase flows describing particle dispersion. ^53^ In our trials, the particles were generated at a steady rate for 60 sec, comparable to Crawford et al., and we also observed highly nonuniform plumes during the aerosolization. Due to the negative pressure ICU’s high air mixing rate, particle concentration was at a well-mixed state ∼30 sec after the aerosolization was stopped.

In summary, these CFD studies could benefit from more robust boundary conditions and spatiotemporal aerosol distribution that could be used for validation. Longer-term simulations that could be used to calculate the decay rate and quantify the aerosol’s nonuniformity could benefit stakeholders.

## 4 Conclusions

These experiments provide the baseline for evaluating the persistence and fate of aerosols in both ICUs and non-ICU clinical spaces. The negative-pressure rooms effectively eliminated aerosol leaks from ICUs; however, positive- and neutral-pressure rooms allowed up to 6 % and 19 % of aerosol escape, respectively. This suggests that closing the door could prevent up to 13% of aerosol leakage, protecting healthcare workers and patients outside an ICU. The nursing stations 10-20 feet outside each ICU consistently did not show PM spikes from the aerosol generation in the ICU. Negative-pressure ICUs clear aerosols to background levels up to 1.6 - 1.8 x faster than a positive-pressure room. Though the negative pressure rooms were superior for managing the amount of aerosol in a room and controlling leakage, the presence of an antechamber also reduced the number of aerosols leaving the room. From the perspective of public health, our measurements indicate that the potentially infectious particles escaping ICU or another pressurized room environment, such as OR, pose a risk for medical personnel and patients in the vicinity; and vice versa, the aerosol from outside the ICU can be introduced into the room from outside into the negative pressure operation.

Using the sensor network measurements informed the development of a reduced-order zonal model for aerosol distribution in ICUs under positive, neutral, and negative pressure. Zonal assignment from an unsupervised learning algorithm provided similar results to the zones found by dividing the room into zones spatially, demonstrating that this methodology is generalizable and expandable to other hospital environments (operating rooms, emergency departments, etc.) or non-healthcare areas such as gyms, restaurants, schools, and office buildings. The proposed dispersion and decay rate analysis can be applied to indoor/outdoor aerosol transport systems. The simple empirical relationships are not based on complex CFD models and can be rapidly developed for complex real-world environments without assumptions associated with boundary conditions and modeling approaches. It can benefit exposure risk assessment, HVAC infrastructure optimization, and intervention strategy development.

## Data Availability

All data produced in the present study are available upon reasonable request to the authors.

## IRB Approval

After consultation with our IRB, it was determined that no IRB was necessary as we made no measurements involving humans.

## Data Availability

The datasets used and/or analyzed during the current study available from the corresponding author on reasonable request

## Notes

### Competing Interest Statement

The authors have declared no competing interest.

### Funding Statement

This study was funded by the NRG research group at the University of Washington

### Summary of Updates

Wording and formatting has been updated. CFD information in the Introduction was added as well as Data Availability after the conclusion.

